# An integrated proteogenomic investigation of the human liver uncovers molecular drivers of steatotic liver disease

**DOI:** 10.64898/2026.06.04.26354903

**Authors:** Émilie Gobeil, Jérôme Bourgault, Malie Enault, Valérie Côté, Patricia L. Mitchell, Louis-Jacques Ruel, Arnaud S. Girard, Marie-Claude Vohl, Benoit J. Arsenault

**Author notes:** <u>Corresponding author information: Benoit Arsenault</u>, PhD, Centre de recherche de l’Institut universitaire de cardiologie et de pneumologie de Québec – Université Laval, Y-3106, Pavillon Marguerite D’Youville, 2725 chemin Ste-Foy, Québec (QC), Canada G1V 4G5.

## Abstract

Metabolic dysfunction-associated steatotic liver disease (MASLD) is rapidly increasing worldwide, yet effective targeted therapies remain limited. To better understand the molecular mechanisms underlying MASLD, we performed an integrated proteogenomic analysis of human liver tissue. Using mass spectrometry, we quantified 2,744 proteins in 504 liver biopsies from the Quebec Obesity Biobank and examined changes across disease stages. To investigate causality, we integrated liver proteomics with RNA sequencing and genome-wide genotyping to map thousands of protein quantitative trait loci (pQTLs) and expression quantitative trait loci (eQTLs). These molecular data were combined with summary statistics from a meta-analysis of genome-wide association studies including 16,532 MASLD cases and 1,240,188 controls. Mendelian randomization and genetic colocalization analyses revealed that most proteins differentially expressed across MASLD stages were not causally implicated in disease risk, whereas several genetically predicted liver proteins showed evidence of causal effects. Among these, higher hepatic levels of the MTARC1 protein were causally associated with MASLD and hepatic fat accumulation. Phenome-wide analyses suggested that MTARC1 inhibition may reduce the risk of cirrhosis, hepatocellular carcinoma, and cholelithiasis while improving lipid profiles. Notably, the causal MTARC1 variant influenced liver protein levels but not gene expression. Genetic analyses also identified ERLIN1 and HSD17B13 as potential therapeutic targets. In contrast, eQTLs and pQTLs at other loci such as *GCKR* showed opposite effects on MASLD risk. These findings highlight the importance of integrating tissue proteomics with human genetics to distinguish biomarkers from causal drivers and to identify promising therapeutic targets for MASLD.

**Graphical abstract:** Created in BioRender. Gobeil, É. (2026) https://BioRender.com/ftsl0yd

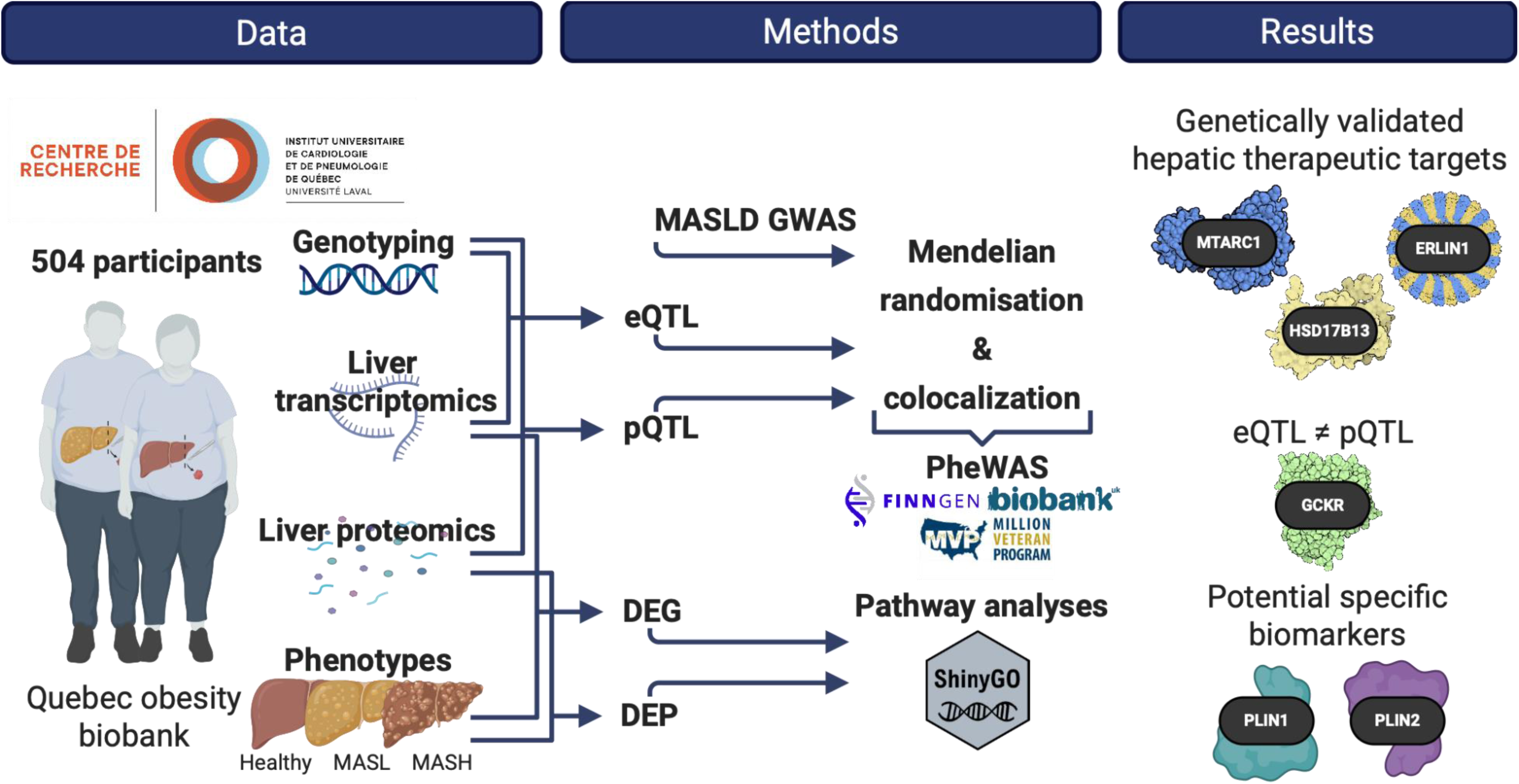

**In brief:** Integrating liver proteomics, transcriptomics, and human genetics in 504 biopsies, this study shows that most proteins altered in MASLD are biomarkers rather than causal drivers. Genetic evidence implicates MTARC1, ERLIN1, and HSD17B13 as potential therapeutic targets and highlights the importance of protein-level regulation in liver disease.

**Key points:** Integrated proteomics, transcriptomics, and human genetics in 504 liver biopsies identified proteins and pathways dysregulated across MASLD stages.

Genetic analyses revealed that most differentially expressed liver proteins represent disease biomarkers rather than causal drivers of MASLD.

Proteogenomic integration identified MTARC1, ERLIN1, and HSD17B13 as potential causal proteins and therapeutic targets, and highlighted discordant effects of eQTLs and pQTLs at loci such as *GCKR*.

## Introduction

Metabolic dysfunction–associated steatotic liver disease (MASLD) affects up to one in three adults worldwide and represents a growing public health burden [1, 2]. Despite its high prevalence and clinical consequences, effective targeted therapies remain limited, in part due to an incomplete understanding of its underlying biology. Large-scale genome-wide association studies (GWAS) have identified numerous loci associated with MASLD, providing important insights into its genetic architecture and highlighting potential therapeutic targets [3–5]. However, translating these findings into biological mechanisms remains challenging. Most associated variants lie in non-coding regions and are thought to exert regulatory effects, complicating the identification of causal genes and pathways [6, 7]. In previous work, we generated liver transcriptomic and genetic data to map expression quantitative trait loci (eQTLs), which enabled the identification of novel protein-coding genes and long non-coding RNAs, including TRIB1AL, as candidate drivers of MASLD [8]. However, transcriptomic studies alone do not capture downstream molecular processes, such as changes in protein abundance, which are more proximal to disease pathophysiology [9, 10].

To address these limitations, integrating genetic data with molecular profiling of disease-relevant tissues offers a powerful approach to identify causal mechanisms [11, 12]. In particular, the study of protein quantitative trait loci (pQTLs) enables the investigation of how genetic variation influences protein abundance, a molecular layer more proximal to cellular function and disease processes than gene expression alone [13, 14]. In this study, we generated a large-scale proteogenomic dataset from human liver biopsies obtained from the Quebec Obesity Biobank, including quantitative proteomics, RNA sequencing, and genome-wide genotyping. Leveraging these data, we mapped liver pQTLs and integrated them with large-scale genetic association data to characterize molecular alterations across the MASLD spectrum, distinguish disease-associated biomarkers from causal drivers using Mendelian randomization and colocalization analyses, and identify and prioritize potential therapeutic targets.

## Materials and methods

### Study design and proteogenomic data acquisition

Patients undergoing bariatric surgery at the Quebec Heart and Lung Institute (*Institut universitaire de cardiologie et de pneumologie de Québec* [IUCPQ]) provided informed consent to participate to the institutional biobank (Quebec Obesity Biobank). Clinical information at the time of surgery, including sex, age, anthropometric measurements, medication use, medical history, comorbidities, and glycemic, lipoprotein and liver enzyme profiles, were available for all patients. Detailed methods of DNA and RNA extraction, genome-wide genotyping, RNA sequencing, transcript expression quantification, and mapping of eQTL has been previously described [8]. A total of 504 samples passed genotyping and RNA sequencing quality controls. Briefly, liver samples were obtained by incisional biopsy of left lobe and were not cauterized. The grading and staging of histological lesions have been carried out according to the protocol of Brunt et al. [15, 16] by pathologists who were blind to the objectives of the study. The liver sample procedure and position of the liver sample are standardized among all surgeons. Liver concentrations of 2744 proteins were measured in 460 liver samples from the same cohort by LC-MS/MS-DIA. Proteins were extracted from liver tissue by sonication in lysis buffer (50 mM ammonium bicarbonate, 0.5% sodium deoxycholate, 50 mM DTT, protease inhibitors). After centrifugation (10,000 × g, 15 min, 4°C), supernatants were acetone precipitated overnight at - 30°C, resuspended in ammonium bicarbonate with 1% sodium deoxycholate, and quantified by Bradford assay. Five micrograms of protein were denatured, reduced, alkylated, and digested overnight with trypsin (Promega). Digestion was quenched with formic acid, sodium deoxycholate was removed by centrifugation, and peptides were dried and resuspended in 0.1% formic acid. Five hundred nanograms of peptides were loaded onto Evotips and analyzed by LC-MS/MS using an Evosep One system coupled to an Orbitrap Exploris 480. Peptides were separated on an 8 cm capillary column using a 21 min gradient (60 samples/day). DIA data were acquired over 350-875 m/z using 35 windows, with MS1 resolution of 120,000 and DIA resolution of 15,000. Raw data were processed with DIA NN v1.8.1 using a predicted UniProt human spectral library. Trypsin/P specificity with up to two missed cleavages was allowed; methionine oxidation and N terminal acetylation were variable modifications, and cysteine carbamidomethylation was fixed. Quantification was limited to peptides of 7-30 amino acids and charge states 2+-5+, with Match Between Runs enabled. Protein intensities were normalized using MaxLFQ, and proteins identified with ≥2 peptides and ≤30% imputed values were retained. PQTL analyses were performed by testing the association between protein abundances and genetic variants. Protein intensities were inverse rank normalized prior to analysis.

Covariates for pQTL mapping included sex, age, and the top 10 genotyping principal components. A set of covariates identified using the Probabilistic Estimation of Expression Residuals (PEER) method [17], calculated for the normalized protein abundances matrices, was also used. For pQTL analyses, protein values were adjusted for 60 PEER factors to improve the number of eProteins discovered. Next, tensorQTL was used for pQTL quantification as described by the developers [18]. Only single-nucleotide variations (SNVs) with a minor allele frequency greater than 0.01 were considered for pQTL mapping. Associations were computed separately for each protein–variant pair, and genome wide significance was defined using a false discovery rate (FDR) of 0.05. Variants within ±1 Mb of the transcription start site of the encoding gene were classified as cis pQTLs.

### Differential expression analysis of hepatic proteins

Differential expression analyses were performed to compare hepatic protein concentrations in two comparisons: first, between participants without MASLD (n = 96) and those with MASLD (n = 408; MASL n = 254 and MASH n = 165), and second, between participants with MASL (n = 254) and those with MASH (n = 165). Hepatic protein concentration refers to a relative concentration determined by mass spectrometry intensity, normalized using rank-based inverse normal transformation as previously described [8]. The DEqMS statistical method, implemented via the R package of the same name, was applied to 2744 proteins with measured relative concentrations. This method incorporates protein-specific measurement uncertainty and employs empirical Bayes variance moderation to improve statistical power and control false-positive findings [19]. Statistical significance was determined using a FDR-adjusted p-value < 0.05. For visualization and interpretation, proteins were further filtered based on effect size. In the comparison between participants without MASLD and those with MASLD, proteins with an absolute log□ fold change (|log□FC|) > 0.5 were considered to exhibit biologically meaningful differences. In the second analysis comparing participants with MASL and MASH, a lower effect size threshold was applied, and proteins with an absolute log fold change (|log□FC|) > 0.2 were considered to display differential relative abundance. The selected fold change thresholds are consistent with prior quantitative proteomics studies, which emphasize that effect size cutoffs should be interpreted in the context of measurement technology and biological contrast. In particular, proteomics analyses commonly apply moderate thresholds (|log□FC| ≈□0.5) for case-control comparisons to highlight robust differences, while permitting lower cutoffs (≈ 0.2-0.3) when assessing disease progression, where fold change compression and subtler but biologically relevant effects are expected, provided strict false discovery rate control is maintained [20, 21].

### Differential gene expression analysis

In parallel with the protein analysis, transcriptomic analyses were conducted to evaluate differences in gene expression levels between participants without MASLD (n = 96), and with MASLD (MASLD n = 408; MASL n = 247 and MASH n = 161). Read counts and TPM values were obtained with RNA-SeQC v2.4.2 [22]. Differential expressed gene (DEG) analysis was performed using DESeq2, a Bioconductor package in R, designed for count-based RNA sequencing data. DESeq2 models gene-level read counts using generalized linear models based on the negative binomial distribution and applies shrinkage estimation for dispersion and fold changes [23]. P-values were adjusted for multiple testing using the Benjamini–Hochberg FDR correction, and genes with an FDR-adjusted p-value < 0.05 were considered statistically significant. However, only genes associated with the proteins of interest were compared at this stage.

### Mendelian randomization analyses

Mendelian randomization (MR) analyses were conducted to evaluate the presence of a causal link between liver protein levels and MASLD. For each protein, two types of exposures were analyzed independently: genetic variants modulating transcript expression (eQTLs) and those modulating protein expression (pQTLs). These were previously assessed in the Quebec Obesity Biobank cohort and described by Gobeil et al., 2025 [8]. As outcome data, primary MR analyses used a genome-wide association study (GWAS) comprising six cohorts and including 16,532 MASLD cases and 1,240,188 controls [3, 5]. For eQTLs and pQTLs, the instrumental variables (genetic variants; SNPs) were filtered using a p-value threshold of ≤ 5×10□□to retain only those significantly associated with hepatic gene expression or protein levels. Additionally, eQTL and pQTL SNPs were filtered to exclude those in linkage disequilibrium via clumping (r2≥0.01), prior to MR analyses. After filtering, only one SNP remained eligible for MR in most exposures, so the Wald estimator was used. Analyses were conducted using the R package TwoSampleMR v.0.5.6 [24]. A p-value ≤ 0.05 was required to consider a causal relationship between exposure and outcome, and to identify the proteins of interest as potential therapeutic targets. For MTARC1-specific analyses, MR was performed using rs2642438 as the instrumental variable, with effect estimates scaled to represent the change in each outcome per 1-standard deviation (SD) decrease in genetically predicted liver MTARC1 protein levels. Summary-level data for the outcomes were obtained from large-scale GWAS conducted in populations of predominantly European ancestry. Liver fat data were derived from Sveinbjornsson et al. [3], including 9,491 cases and 876,210 controls from deCODE genetics, UK Biobank (UKBB), FinnGen, and U.S.-based cohorts. MASLD GWAS data were identical to those used in the primary MR analysis. Liver cirrhosis data were obtained from Ghouse et al. [25], including 15,225 cases and 1,564,786 controls from cohorts across the United States, Finland, Denmark, the United Kingdom, Iceland, Germany, and Estonia, incorporating data from All of Us and the Million Veteran Program (MVP). Hepatocellular carcinoma GWAS data were obtained from Ghouse et al. [26], including 3,748 cases and 1,861,536 controls from similar cohorts.

### Pairwise conditional and colocalization analysis (PWCoCo)

PWCoCo was used to perform conditional analyses within each locus to identify conditionally independent signals for MASLD and the corresponding eQTL or pQTL genomic region [27]. Colocalization analyses were then conducted for each pair of conditionally independent signals between the two traits (eQTLs or pQTLs and MASLD) using summary-level data. This approach ensures that the stringent single-variant assumption holds for each colocalization test. Briefly, this framework integrates approximate conditional analyses (as implemented in GCTA-COJO [28]), which systematically conditions on each of the association signals within a genomic region and applies pairwise Bayesian colocalization analyses for each pair of independent signals (as implemented in the coloc R package [29]).

### Fine-mapping and pathway analysis

To prioritize candidate genes underlying multi-omics associations with MASLD, fine-mapping and pathway enrichment analyses were performed. Genes identified through differential expression, proteomic analyses, and genetic association or Mendelian randomization approaches were compiled into a unified gene set for downstream functional interpretation. Pathway enrichment analysis was conducted using Kyoto Encyclopedia of Genes and Genomes (KEGG) pathways, a curated database that maps genes to known biological pathways involved in metabolism, signaling, cellular processes, and disease mechanisms [30]. KEGG analysis was used to identify biological pathways significantly overrepresented among the candidate genes, providing insight into shared molecular functions and disease-relevant processes. Functional enrichment and pathway analyses were implemented using ShinyGO (version 0.85), an interactive web-based tool for gene set enrichment analysis [31]. ShinyGO was used to test for overrepresentation of KEGG pathways among the input gene list relative to a genome-wide background. Enrichment p-values were adjusted for multiple testing using the Benjamini–Hochberg FDR correction, and pathways with an FDR-adjusted p-value < 0.05 were considered statistically significant. Visualization outputs from ShinyGO were used to summarize enriched pathways and gene–pathway relationships.

### Data availability

All GWAS summary statistics used in this study are publicly available. The GWAS for MASLD was the same as that used in our previous study [8]. The GWAS for liver fat was downloaded from deCODE genetics consortium page: https://www.decode.com/summarydata. GWAS data for cirrhosis of liver and hepatocellular carcinoma were obtained from the GWAS Catalog under accession numbers GCST90319877 and GCST90809296, respectively. PheWAS data were retrieved on the FinnGen portal: https://public-mvp-ukbb.finngen.fi/. All data required to evaluate or replicate this work are provided in the Supplementary Table 1.

### Disclosure on the use of generative AI

Portions of the manuscript were revised for clarity and language using an AI-based language model (M365 Copilot, GPT-5–based). The authors reviewed and edited all outputs and take full responsibility for the final content.

### Ethical approval

The study was conducted in accordance with the Declaration of Helsinki and approved by the Institutional Review Board (Ethics Committee) of Institut universitaire de cardiologie et de pneumologie de Québec-Université Laval (IUCPQ-UL) (approval number 2021-3656; date of approval: 17 June 2021). For the other publicly available GWAS summary statistics, all participants provided informed consent, and study protocols were approved by their respective local ethical committee.

## Results

### Differentially expressed proteins associated with steatohepatitis and fibrosis

To characterize both disease presence and progression, hepatic concentrations of 2,744 proteins were analyzed using two comparison schemes: (i) no MASLD (control) versus MASL or MASH, and (ii) MASL versus MASH. After p-value adjustment, 194 proteins showed significantly higher levels in MASLD condition compared with control condition, whereas 38 proteins showed lower levels (Figure 1A, and Supplementary Table 1). These 194 proteins were enriched in multiple KEGG metabolic pathways like: other glycan degradation, collecting duct acid, complement and coagulation cascades, PPAR signaling pathway, cholesterol metabolism, lysosome, carbon metabolism, FcγR-mediated phagocytosis, apoptosis, MTOR signaling pathway, endocytosis, and regulation of actin cytoskeleton (Figure 1B right panel). For proteins with lower abundance in MASLD condition, they are involved also in metabolic pathways such as fatty acid degradation, retinol metabolism, pyruvate metabolism, biosynthesis and metabolism of amino acids, and pentose and glucuronate interconversions (Figure 1B left panel).

**Figure 1.**
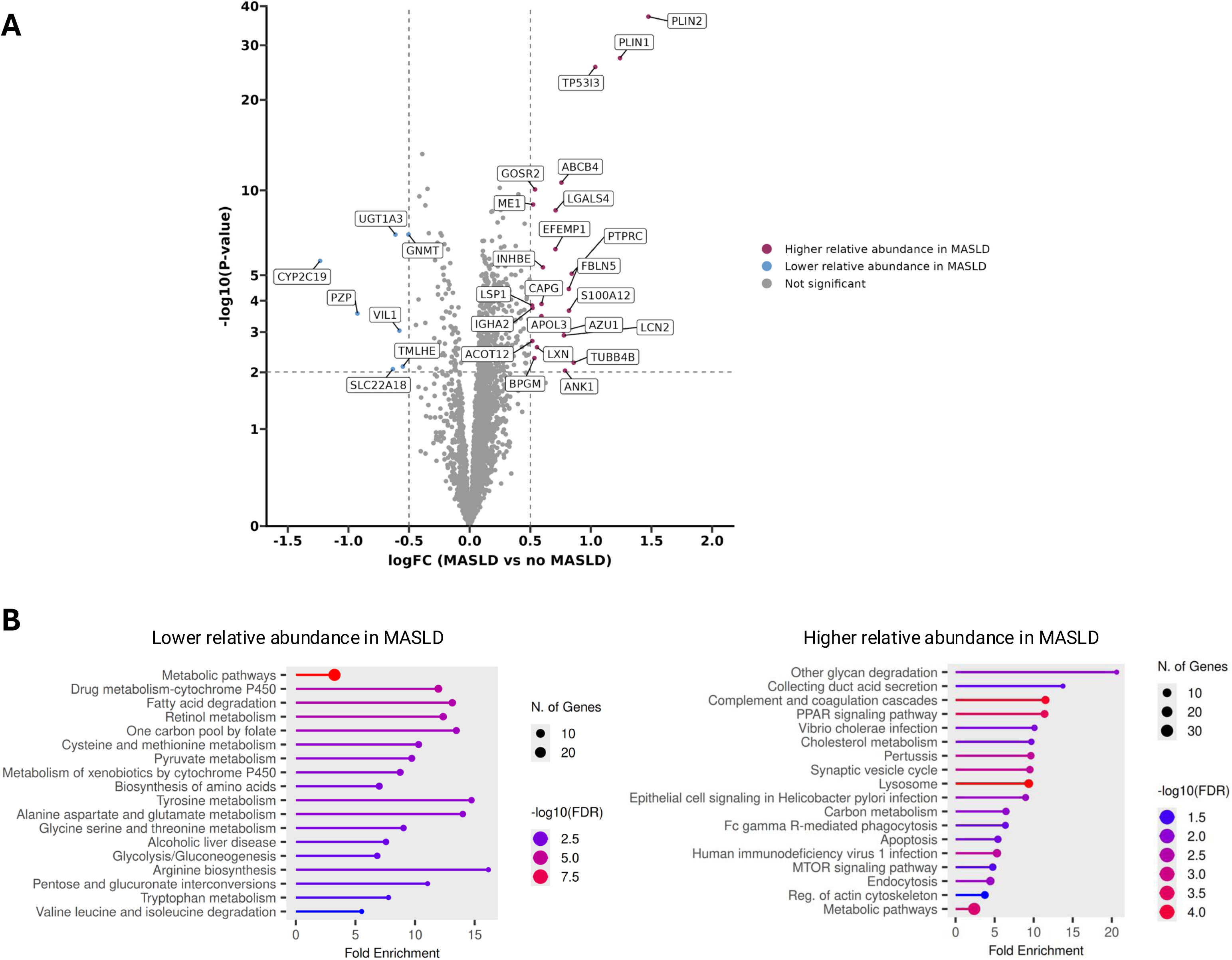
Differentially expressed proteins in participants with steatosis (MASLD; n=357) compared to participants without steatosis (no MASLD; n=106). **(A)** shows a volcano plot with the results of the differential expressed protein (DEP) analysis for the Quebec Obesity Biobank cohort. **(B)** presents pathways groupments obtained with shinyGO v0.85 for key gene function enrichment analysis with KEGG for downregulated proteins (left panel) and for overexpressed proteins (right panel). A positive fold changes correspond to proteins more abundant in participants without MASLD, and vice versa.

In Figure 1A, positive fold changes correspond to proteins more abundant in participants without MASLD, and vice versa. It is observed that perilipins 1 and 2 (PLIN1 and PLIN2) show the greatest concentration differences between the two subgroups. Participants with MASLD have 24% higher PLIN1 concentration (p = 7×10 ²) and 47% higher PLIN2 concentration (p = 2×10 ³) compared to participants without MASLD (Supplementary Table 1). Both proteins have a FDR ≤ 0.01, which reinforces the significance of the results.

To observe the proteins that may be implicated in the progression of steatosis to fibrosis, liver proteins levels were compared between MASL and MASH conditions. After p-value adjustment, 44 proteins showed significantly higher abundance in MASH compared with controls, whereas 8 proteins showed lower abundance (Figure 2A, and Supplementary Table 1). Proteins with higher levels in MASH were primarily enriched in pathways related to neuroactive ligand-receptor interaction, IL-17 signaling pathway, motor proteins, regulation of actin cytoskeleton, and tight junctions (Figure 2B right panel). Among the 8 proteins with lower abundance, only two (ADH1B and ACADSB) formed a network in KEGG pathway enrichment analysis of fatty acid degradation (Figure 2B left panel).

**Figure 2.**
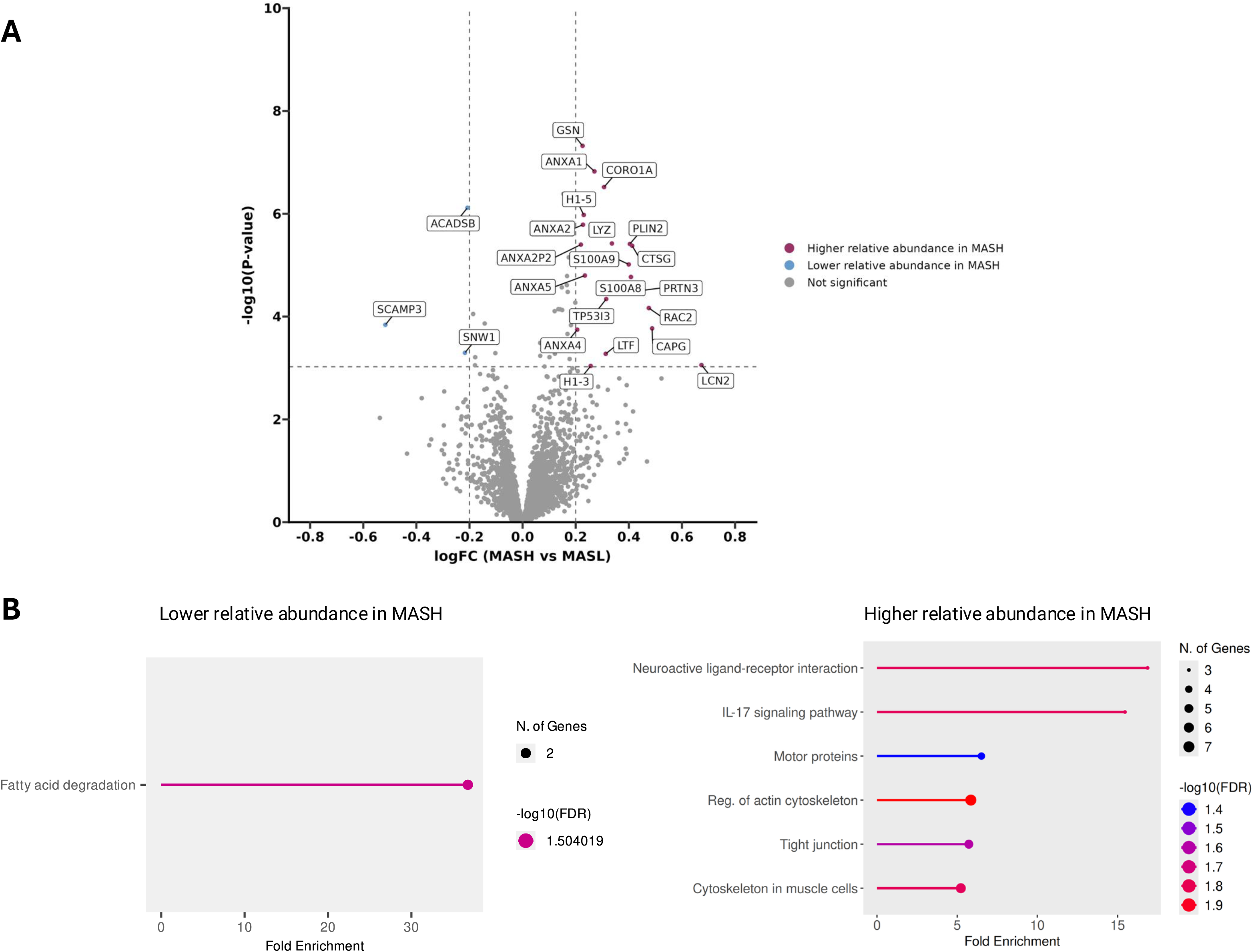
Differentially expressed proteins in participants with fibrosis compared to participants without fibrosis. **(A)** shows a volcano plot with the results of the differential expressed protein (DEP) analysis for the Quebec Obesity Biobank cohort. **(B)** presents pathways groupments obtained with shinyGO v0.85 for key gene function enrichment analysis with Kyoto Encyclopedia of Genes and Genomes (KEGG) for significatively downregulated proteins (left panel) and for significatively overexpressed proteins (right panel).

### Identification of liver protein quantitative trait loci associated with MASLD

To identify liver-expressed proteins that are potentially causally associated with MASLD, we performed a combination of MR and genetic colocalization analyses using liver pQTLs derived from the Quebec Obesity Biobank. Among 2744 distinct liver protein levels measured, at least one genome-wide significant pQTL association (p ≤ 5e-08) was identified for 643 proteins, which were carried forward for MR analyses (Supplementary Table 1). MR results are summarized in Figure 3, which depicts the effect estimates of genetically predicted liver protein levels on MASLD risk. Several proteins reached statistical significance after correction for multiple testing, showing both positive and negative associations with disease risk. To distinguish potentially causal effects from associations driven by linkage disequilibrium (LD), genetic colocalization analyses were performed for MR significant protein–MASLD associations. This analysis identified ERLIN1, MTARC1, GCKR, LGALS1, and MTTP as proteins influencing MASLD risk (p_FDR_ ≤ 0.1) and exhibiting strong evidence of shared genetic signals with MASLD (posterior probability of colocalization, PP_H4_ ≥ 80%; Table 1). In contrast, most of MR supported associations (9 of 14) did not show strong evidence of colocalization (PP_H4_ < 80%; Table 1), highlighting the potential for confounding by LD and underscoring the importance of colocalization analyses in protein based MR studies. Using MR instruments derived from cis pQTLs, MTARC1 showed a strong positive association with MASLD risk (β□= 0.231, SE = 0.028, P = 1.52 × 10 ¹), with complete evidence of colocalization (PP_H4_ = 1.00). Similarly, GCKR demonstrated a strong inverse association with MASLD using pQTL based MR (β = -0.111, SE = 0.011, p = 3.24 × 10 ² ; PP_H4_ = 1.00). For both proteins, MR analyses using eQTL instruments showed little to no evidence of colocalization (*MTARC1* PP_H4_ = 0.003; *GCKR* PP_H4_ = 6.8 × 10), despite a strong MR eQTL association for *GCKR* (β = 0.402, SE = 0.058, p = 4.19 × 10 ¹²) and the absence of a significant pQTL for MTARC1 to enable MR pQTL analysis, indicating that genetic effects on MASLD are predominantly mediated at the protein level. For its part, LGALS1 exhibited consistent MR effects at both molecular layers. MR eQTL analysis showed a negative association with MASLD risk (β = -0.065, SE = 0.017, p = 1.57 × 10 ; PP_H4_ = 0.87), which was concordant with the MR pQTL estimate (β = -0.075, SE = 0.020, p = 1.57 × 10 ; PP_H4_ = 0.87), both driven by the same lead variant (rs7291867), supporting coordinated genetic regulation across transcript and protein levels. ERLIN1 demonstrated a significant MR pQTL association with MASLD (β = 0.187, SE = 0.039, p = 1.84 × 10) and strong colocalization (PP_H40_= 0.98), whereas the MR eQTL estimate was weaker and non significant (β = -0.080, SE = 0.066, P = 0.22). *MTTP* showed significant MR eQTL (β = -0.501, SE = 0.087, p = 1.03 × 10) and MR pQTL (β = -0.218, SE = 0.069, p = 0.001) associations with MASLD, with strong evidence of colocalization at the protein level (PP_H4_ = 0.84). Together, integration of MR, pQTL, eQTL, and colocalization analyses prioritized ERLIN1, MTARC1, GCKR, LGALS1, and MTTP as liver expressed proteins robustly associated with MASLD risk, supported by shared genetic signals at the protein level and, for selected targets, coordinated regulation at the transcript level.

**Figure 3.**
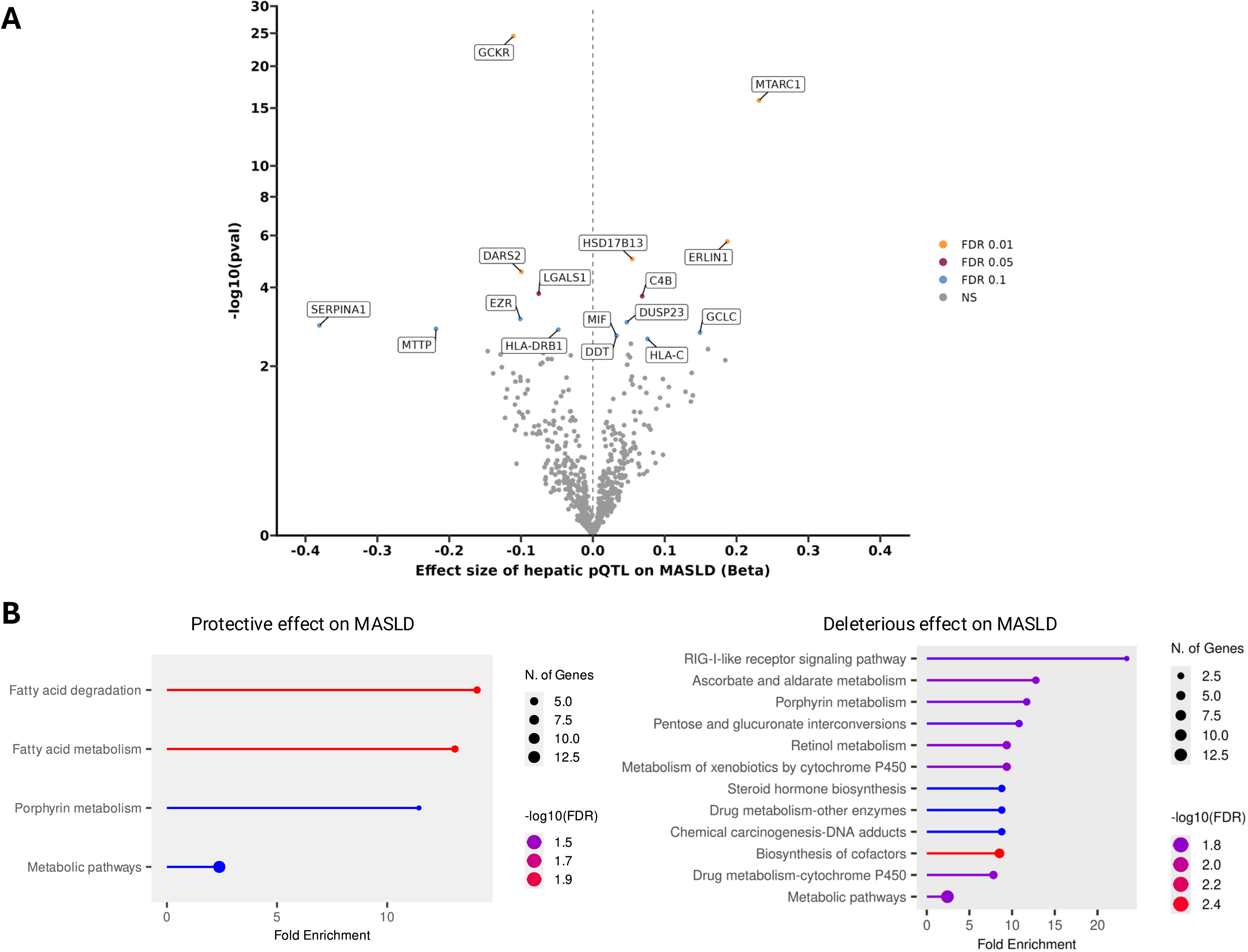
Mendelian randomization analysis of genetically predicted liver expressed proteins and MASLD risk. Volcano plot of MR results between liver pQTLs and MASLD. Each point represents a protein instrumented by liver pQTLs. The x axis indicates the MR effect estimate (β) of protein levels on MASLD, and the y axis shows the −log (P value). Proteins are colored according to FDR, and selected proteins with significant or notable associations are labeled.

**Table 1.** Mendelian randomization and colocalization results of hepatic pQTL impacting MASLD and the results of the equivalent hepatic eQTL on MASLD.

|  | MR-eQTL |  |  |  |  | MR-pQTL |  |  |  |  |
| --- | --- | --- | --- | --- | --- | --- | --- | --- | --- | --- |
| Protein | rsID | Beta | SE | Pval | coloc<br>ppH4 | rsID | Beta | SE | Pval | coloc<br>ppH4 |
| **GCKR** | rs1260326 | 0.402 | 0.058 | 4.19E-12 | 6.79E-07 | rs1260326 | -0.111 | 0.011 | 3.24E-25 | 1.00 |
| **MTARC1** |  |  |  |  | 0.003 | rs2642438 | 0.231 | 0.028 | 1.52E-16 | 1.00 |
| **ERLIN1** | rs7477551 | -0.080 | 0.066 | 0.222 | 0.005 | rs1408579 | 0.187 | 0.039 | 1.84E-06 | 0.98 |
| **LGALS1** | rs7291867 | -0.065 | 0.017 | 1.57E-04 | 0.866 | rs7291867 | -0.075 | 0.020 | 1.57E-04 | 0.87 |
| **MTTP** | rs7665143 | -0.501 | 0.087 | 1.03E-08 | 0.945 | rs17029137, rs2035816 | -0.218 | 0.069 | 0.001 | 0.84 |
| **HSD17B13** |  |  |  |  | 0.043 | rs6831888 | 0.055 | 0.012 | 9.69E-06 | 0.79 |
| **DUSP23** | rs3806187 | 0.002 | 0.050 | 0.970 | 0.006 | rs1129923 | 0.047 | 0.014 | 0.001 | 0.63 |
| **SERPINA1** |  |  |  |  | 0.029 | rs17580 | -0.381 | 0.118 | 0.001 | 0.56 |
| **EZR** | rs901367 | -0.079 | 0.024 | 0.001 | 0.482 | rs901367 | -0.101 | 0.030 | 0.001 | 0.44 |
| **GCLC** |  |  |  |  | 0.086 | rs545751 | 0.149 | 0.048 | 0.002 | 0.29 |
| **MIF** | rs5760119 | 0.028 | 0.010 | 0.004 | 0.367 | rs5751775 | 0.033 | 0.011 | 0.002 | 0.29 |
| **DDT** | rs5760119 | 0.031 | 0.010 | 0.002 | 0.369 | rs5751775 | 0.033 | 0.011 | 0.002 | 0.29 |
| **C4B** | rs443198 | 0.005 | 0.046 | 0.909 | 0.008 | rs3130614, rs3957147 | 0.069 | 0.01 | 1.88E-04 | 0.03 |
|  |  |  |  |  |  |  |  | 8 |  |  |
| **DARS2** | rs72709386 | 0.048 | 0.068 | 0.481 | 0.007 | rs141915768,<br>rs144366549 | -<br>0.099 | 0.02<br>4 | 2.94E-05 | 0.01 |

### Fine-mapping and assessment of convergence and divergence between omics analyses

To distinguish proteins that are likely causal for MASLD from those that may represent downstream consequences of the disease, we assessed convergence and divergence across five complementary omics analyses (Supplementary Table 2): differential gene expression (DEG), differentially expressed proteins (DEP), GWAS-significant single-nucleotide polymorphisms (SNPs), Mendelian randomization using expression quantitative trait loci (MR-eQTL), and Mendelian randomization using protein quantitative trait loci (MR-pQTL). The UpSet plot (Figure 4) summarizes the overlap among 668 significant hits identified across these types of analyses. Most significant results were specific to a single analytical layer, with 334 hits unique to DEG and 195 unique to DEP, highlighting substantial modality-specific regulation. A smaller subset of genes and proteins showed evidence of overlap between transcriptomic and proteomic analyses, including 61 hits shared between DEG and DEP, indicating concordant regulation at both the RNA and protein levels. Integration with genetic evidence further reduced the number of convergent candidates. Only a limited number of hits overlapped with GWAS-significant loci and MR-based analyses, underscoring the stringency of causal inference. Notably, *MTTP* emerged as the only gene supported simultaneously by a GWAS-significant SNP and by both MR-eQTL and MR-pQTL analyses, providing consistent genetic evidence for its potential causal protective role in MASLD (Supplementary Table 3). In contrast, *LGALS1* showed significant evidence from differential expression and MR-pQTL analyses but lacked support from GWAS-significant SNPs or MR-eQTLs, suggesting that altered protein abundance of *LGALS1* may reflect downstream effects of disease rather than a primary genetic driver. Additional candidates demonstrated partial convergence across two or three analytical layers, reflecting heterogeneous mechanisms and emphasizing the importance of multi-omics integration to disentangle causality from consequence. Overall, Figure 4 highlights substantial divergence across omics layers, with only a small number of proteins supported by convergent genetic, transcriptomic, and proteomic evidence, thereby prioritizing high-confidence candidates for causal involvement in MASLD pathogenesis.

**Figure 4.**
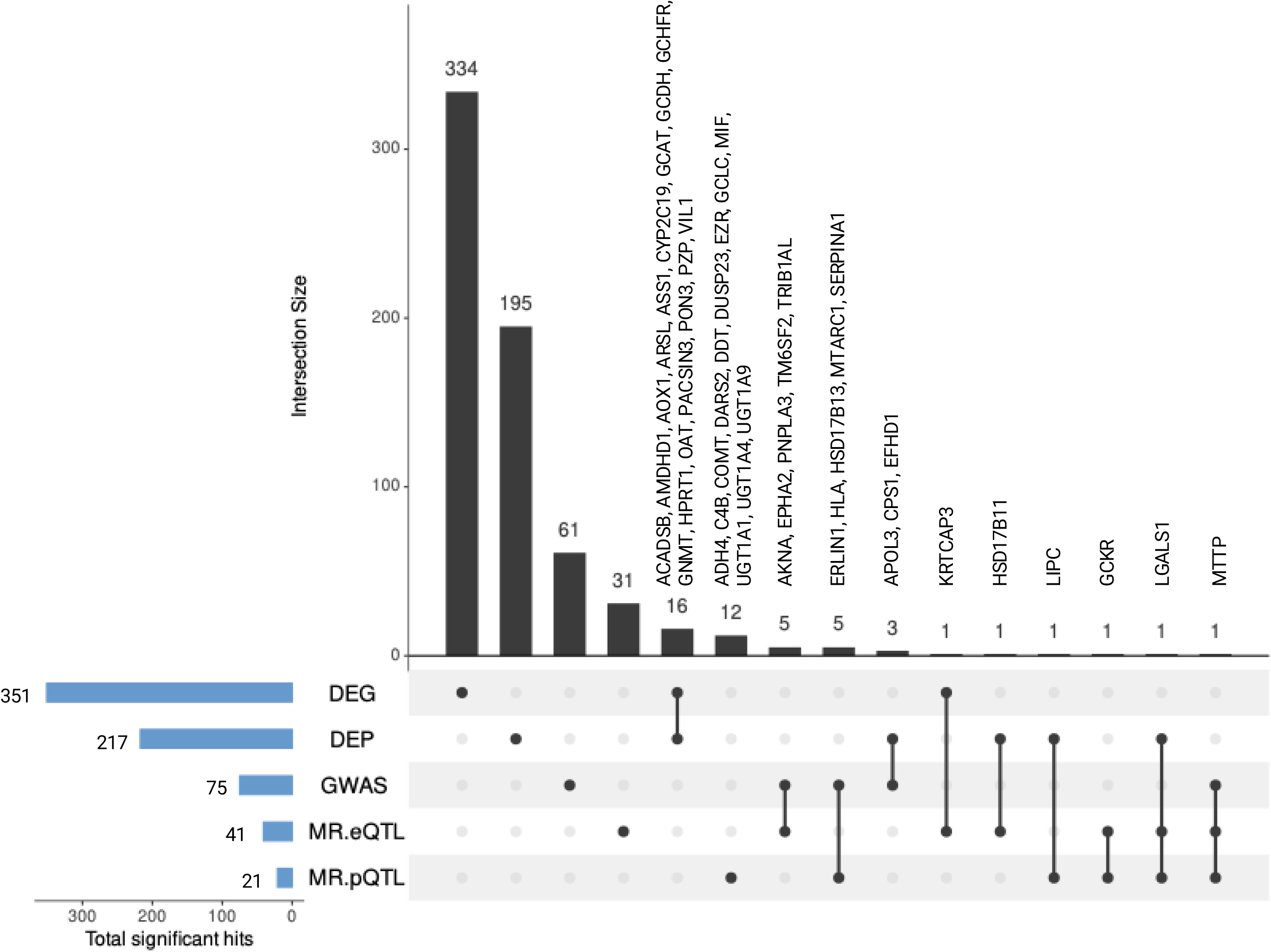
Convergence and divergence of multi-omics analyses associated with MASLD. UpSet plot summarizing the overlap of 668 significant hits identified across five analytical layers: differential gene expression (DEG), differentially expressed proteins (DEP), GWAS significant single nucleotide polymorphisms (GWAS), Mendelian randomization using expression quantitative trait loci (MR eQTL), and Mendelian randomization using protein quantitative trait loci (MR pQTL) as exposures and MASLD GWAS as outcome. Vertical bars represent the size of each intersection, while filled dots and connecting lines indicate the specific combination of analyses contributing to each intersection. Horizontal bars denote the total number of significant hits detected in each individual analysis. Selected genes and proteins with evidence of convergence across multiple omics layers are labeled.

### Effects of genetically predicted reductions in hepatic MTARC1 levels on liver phenotypes

The lead liver pQTL associated with hepatic MTARC1 protein levels and MASLD risk was rs2642438. Given the strong MR and colocalization evidence implicating MTARC1, we further examined the relationship between rs2642438 genotype and hepatic MTARC1 expression and protein levels in the Quebec Obesity Biobank. As shown in Figure 5A, rs2642438 genotype was not associated with differences in hepatic *MTARC1* expression levels. Pairwise comparisons indicated no statistically significant differences in hepatic *MTARC1* expression between genotype groups after adjustment for multiple testing (all adjusted p-value > 0.1). In contrast, rs2642438 genotype was significantly associated with hepatic MTARC1 protein levels (Figure 5B). Mean protein levels increased across genotypes, with lower values in AA homozygotes (n = 39) and in AG heterozygotes (n = 180) than in GG homozygotes (n = 214). Individuals carrying at least one G allele (AG or GG) exhibited significantly higher hepatic MTARC1 protein levels compared with AA homozygotes, with the strongest difference observed between AA and GG genotypes (adjusted p-value = 1.17 × 10 ^4^). Together, these results indicate that rs2642438 is associated with a graded increase in hepatic MTARC1 protein levels that is not fully mirrored at the transcript level, supporting a predominant role for post transcriptional regulation of MTARC1 in the liver. MR analyses revealed that genetically predicted lower liver MTARC1 protein levels were associated with a consistent reduction in risk across the MASLD spectrum (Figure 6). Specifically, a 1-SD decrease in liver MTARC1 protein levels was associated with lower liver fat content (OR 0.88, 95% CI 0.85-0.92; p = 9.39×10) and reduced risk of MASLD (OR 0.79, 95% CI 0.74-0.85; p = 6.10×10 ¹²). Stronger protective effects were observed for more advanced liver disease, including cirrhosis (OR 0.74, 95% CI 0.68-0.80; p = 6.76×10 ¹) and hepatocellular carcinoma (OR 0.69, 95% CI 0.60-0.80; p = 7.08×10). Overall, these findings suggest that reduced MTARC1 protein levels may confer protection against both early and advanced stages of MASLD.

**Figure 5.**
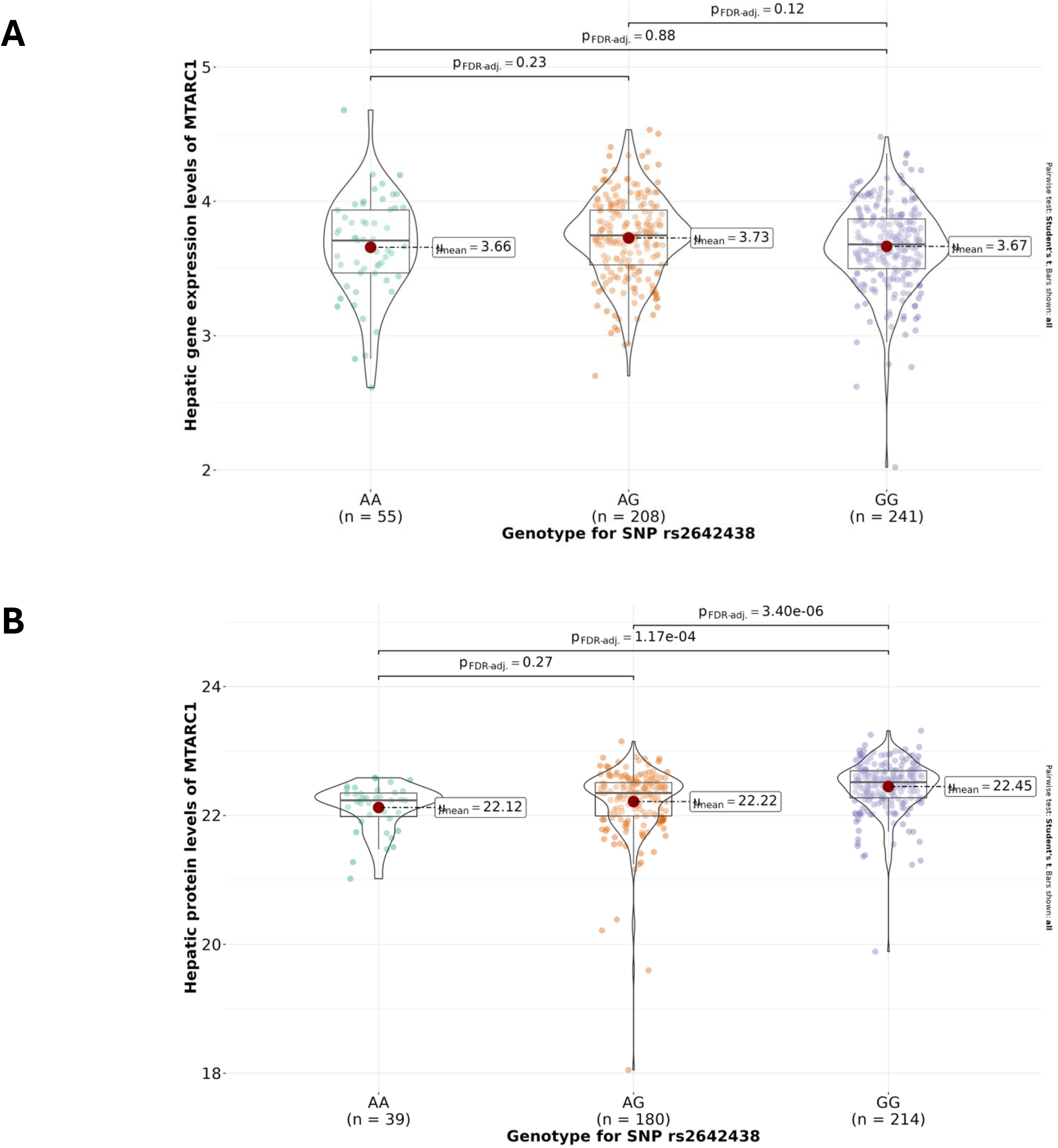
Association between rs2642438 genotype and hepatic MTARC1 expression and protein abundance. **(A)** Hepatic *MTARC1* gene expression levels (transcripts per million; TPM) stratified by rs2642438 genotype (AA, AG, GG). **(B)** Hepatic MTARC1 relative protein levels stratified by rs2642438 genotype. Violin plots depict the distribution of values, with individual data points shown as jittered dots. The central horizontal line within each violin indicates the median, and the boxed area represents the interquartile range. Mean values for each genotype group are indicated by red dots and annotated numerically. Sample sizes for each genotype are shown below the x axis. Pairwise comparisons between genotype groups were performed, with corresponding P values displayed above the plots.

**Figure 6.**
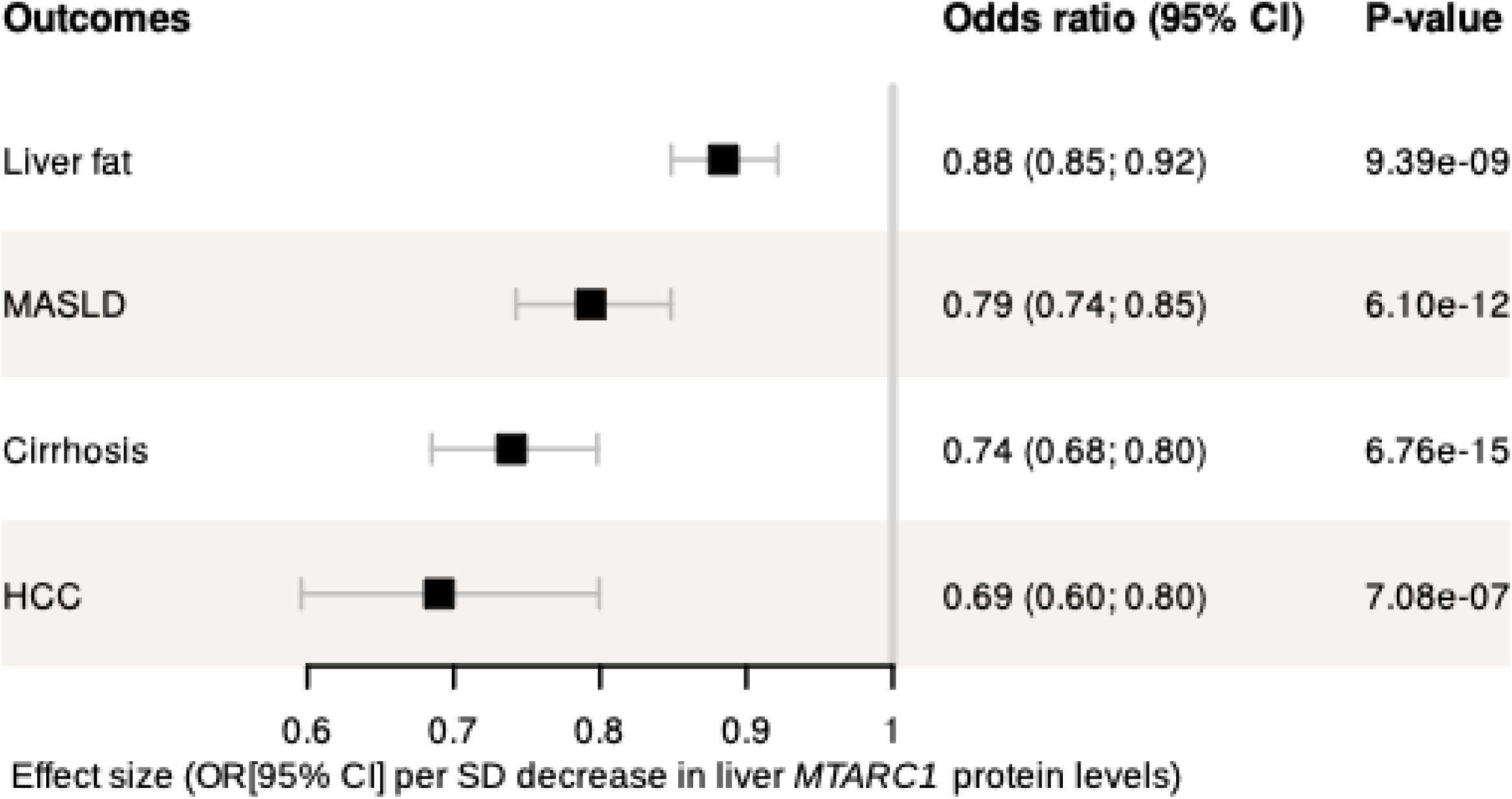
Impact of genetically predicted reductions in liver MTARC1 protein levels on liver diseases. The effects of the strongest SNP associated with the liver MTARC1 protein levels (rs2642438:A) on liver fat, MASLD, cirrhosis and hepatocellular carcinoma were estimated using the Wald ratio in Mendelian randomization analyses. Effect sizes (OR [95% CI]) are presented as the log(OR) change in each outcome per 1-SD decrease in liver MTARC1 protein levels.

### Phenome-wide association study of genetically predicted reductions in hepatic MTARC1 levels

As MTARC1 may represent a therapeutic candidate for the treatment of MASLD, we assessed the potential health benefits or hazards associated with liver MTARC1 inhibition. We look up the phenotypic effects of the rs2642438 variant across 330 traits using meta-analyses of phenome-wide association studies from the Million Veteran Program (n = 449,042), FinnGen (n = 500,349), and the UK Biobank (n = 420,531). Results suggest that MTARC1 inhibition could reduce the risk of cirrhosis (β = -0.08; p = 2.7 × 10 ¹³), hepatocellular carcinoma (β = -0.12, p = 7.9 × 10) and cholelithiasis (β = -0.02, p = 9.3 × 10), while improving the lipid profile (dyslipidaemia: β = -0.02, p = 1.8 × 10 ; hypercholesterolaemia: β = 0.02, p = 4.9 × 10), but potentially increase the risk of idiopathic gout (β = 0.06, p = 4.0 x 10^-5^) or gout (β = 0.02, p = 1.6 × 10^-4^). To a lesser extent, inhibition of MTARC1 was also associated with reduced risk of infective dermatitis (β = -0.03, p = 1.5 × 10 ^4^) and renal failure (β = -0.01, p = 7.8 × 10^-4^) (Figure 7).

**Figure 7.**
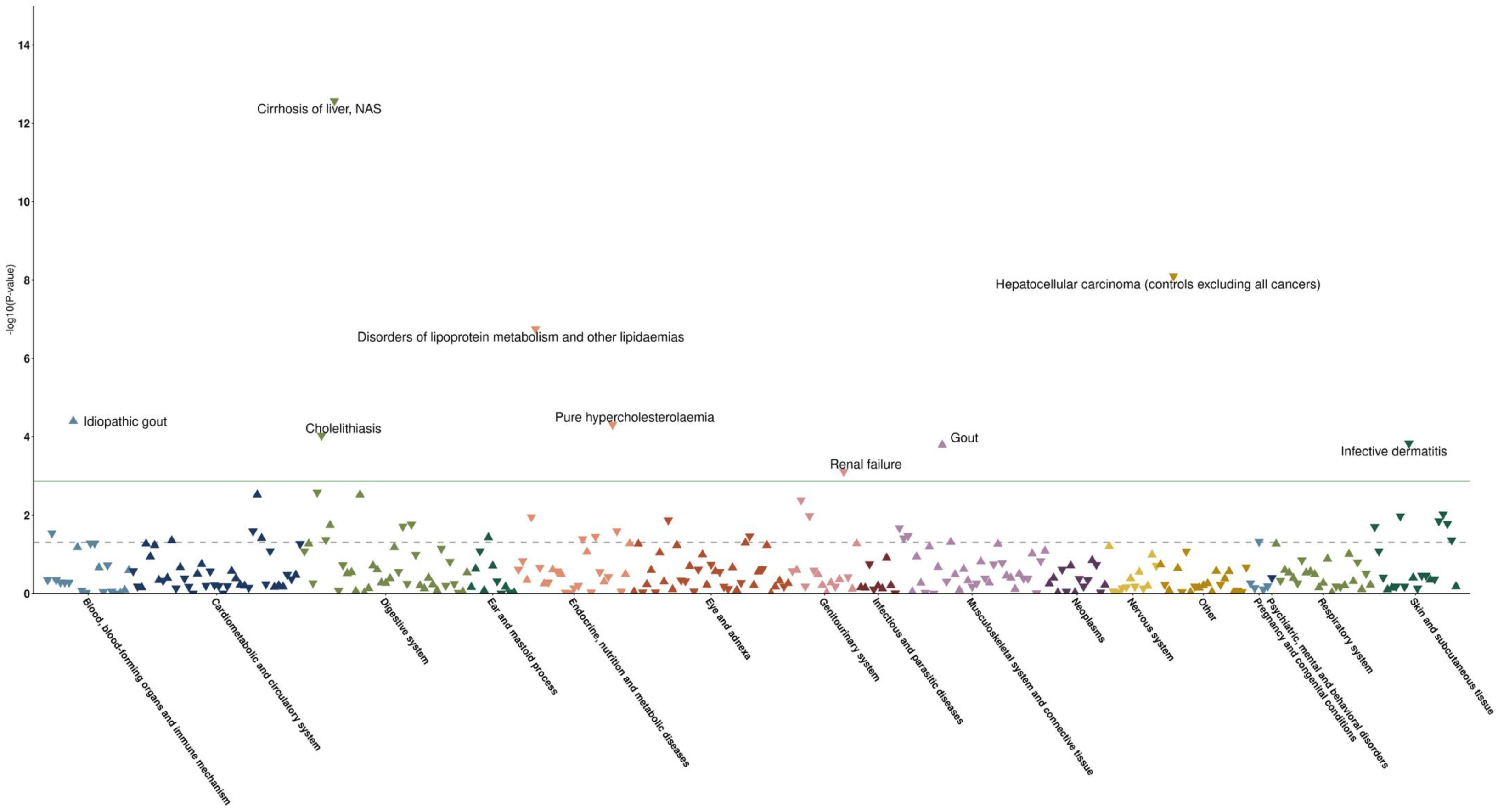
Phenome wide association of rs2642438:A, a genetic proxy for MTARC1 inhibition. PheWAS green horizontal line denotes the significance level after Bonferroni correction for the number of phenotypes testing, conservatively considering each as independent. Phenome wide association study (PheWAS) of the rs2642438:A allele across UK Biobank disease phenotypes. Each triangle represents an individual phecode defined phenotype, grouped along the x axis by disease category and plotted on the y axis according to the strength of association expressed as –log (p value). Triangle orientation indicates direction of effect per rs2642438:A allele (mimicking a MTARC1 inhibition), with upward pointing triangles denoting positive associations (increased disease risk) and downward pointing triangles denoting negative associations (decreased disease risk). Colors correspond to phenotype code categories. The horizontal green line indicates the FDR-adjusted significance threshold (FDR < 0.05), while the dashed grey line denotes nominal significance. Phenotypes exceeding the FDR threshold were considered statistically significant and are annotated.

## Discussion

In this study, we characterized molecular alterations across the spectrum of MASLD and identified proteins that may contribute to disease development. We first compared protein levels among individuals without steatosis (control) to those with steatosis without or with inflammation/fibrosis (MASLD), and after those with steatosis alone (MASL) to those with fibrosis (MASH). These analyses revealed extensive proteomic remodeling in MASLD and additional pathway-level perturbations in MASH, highlighting biological processes that likely underpin disease initiation, progression, and hepatic tissue remodeling. The integration of differentially expressed proteins, pathway enrichment, pQTL mapping, MR, and phenome-wide analyses allowed us to distinguish proteins that likely drive MASLD pathogenesis from those reflecting downstream consequences of MASLD progression.

### Protein levels across the MASLD spectrum

We identified 194 proteins upregulated and 38 downregulated in MASLD compared to controls. Enrichment analyses revealed that the more abundant proteins in MASLD were predominantly involved in metabolic and immunometabolic pathways. Collectively, these enrichments align with the view that early MASLD reflects not only lipid accumulation but also broad changes in intracellular trafficking, innate immune engagement, and stress response pathways. Less abundant proteins in MASLD were mainly enriched in pathways related to hepatic oxidative and intermediary metabolism (i.e., interconnected intracellular pathways governing carbohydrate, lipid, and amino acid metabolism), in line with prior evidence showing that hepatic steatosis is associated with reduced mitochondrial β oxidation, impaired carbohydrate handling, and disruption of central carbon metabolism [32]. In MASH, 44 proteins showed higher levels and only 8 showed lower levels compared to MASL condition. Enriched pathways included IL 17 signaling, neuroactive ligand–receptor interactions, regulation of the actin cytoskeleton, tight junction biology, and motor proteins. All processes aligned with hepatocellular injury, inflammation, immune cell recruitment, and early architectural remodeling that precedes fibrosis [33].

### Genetic analyses candidate proteins with likely causal roles in MASLD

We integrated large-scale liver pQTLs mapping with MR and genetic colocalization. A subset of proteins showed concordant evidence across multiple analytic layers, including GWAS association, significant pQTL-based MR effects, and robust colocalization. These include MTTP, TM6SF2, HSD17B13, MTARC1, ERLIN1 and SERPINA1, which represent particularly compelling candidates because they satisfy stringent causal inference criteria and map to biologically relevant pathways in MASLD. Many of these proteins are involved in hepatic lipid handling and susceptibility to injury. For example, MTTP and TM6SF2 have established roles in VLDL assembly and secretion, connecting genetic variation to hepatic triglyceride retention and steatosis severity. Unfortunately, systemic MTTP inhibition reduces circulating lipids, but it worsens hepatic steatosis, which halted early development of lomitapide like mechanisms for fatty liver [34]. TM6SF2 loss of function variants strongly increase hepatic fat while lowering circulating lipids, creating an unfavorable cardiometabolic trade off [35]. Accordingly, MTTP and TM6SF2 are not pursued to be directly inhibited in MASLD therapeutic strategies.

HSD17B13 and MTARC1 have been implicated in liver injury and disease progression risk in previous human studies, suggesting that variation in hepatocyte stress resilience and metabolic detoxification capacity may shape the trajectory from benign steatosis to inflammatory/fibrotic disease [36–38]. HSD17B13 is the most advanced genetically supported MASLD target in clinical development. Loss of function variants protect against steatohepatitis progression and fibrosis, making enzymatic inhibition attractive [39, 40]. There are three ongoing trials showing active clinical development [41–43]. MTARC1 loss of function variants are associated with protection against MASLD, lower liver enzymes, and reduced liver related mortality without adverse cardiovascular effects. This favorable profile has driven rapid therapeutic translation [44, 45]. Also, preclinical GalNAc siRNA studies show robust reduction in hepatic steatosis across multiple diet induced MASH models [46, 47]. MTARC1 is now widely regarded as a next generation genetic target with high therapeutic index [48]. ERLIN1 plausibly links to ER-associated processes and lipid/sterol homeostasis and has recently emerged as a gain of function protective genetic modifier in MASLD via stabilization of the ERLIN-TM6SF2-APOB complex. Large scale biobank analyses demonstrate that the ERLIN1 p.Ile291Val variant confers reduced MASLD risk, particularly in metabolic and alcohol associated liver disease [49]. Despite compelling human genetics, ERLIN1 targeted therapies remain preclinical. It is currently viewed as a pathway anchor rather than a direct drug target due to its role as a scaffold protein in ER lipid regulation. Ongoing work is focused on identifying druggable downstream nodes, rather than ERLIN1 itself [50]. SERPINA1 is classically linked to alpha 1 antitrypsin deficiency (AATD), it intersects clinically with MASLD via fibrosis risk, proteotoxic stress, and shared metabolic pathways [51, 52]. Although not MASLD specific, SERPINA1 trials inform fibrosis directed and proteostasis based therapeutic strategies applicable to metabolic liver disease [53]. The triangulation of evidence for this group of proteins strengthens the notion that these may not merely represent biomarkers of disease state but could have causal roles in MASLD.

### Liver and phenome-wide effects of lower liver MTARC1 protein levels

MTARC1 was identified as one of the most promising candidate proteins causally linked with MASLD. Fine mapping of pQTLs pinpointed rs2642438 as the top variant regulating liver MTARC1 levels and influencing MASLD susceptibility. Individuals carrying the G allele exhibited significantly higher hepatic MTARC1 concentrations and a higher mean steatosis percentage, consistent with previous studies. Our study also revealed potentially protective effects of MTARC1 on liver cirrhosis and hepatocellular carcinoma. Phenome wide association analysis across 330 traits provided additional insights into the broader physiological implications of altering MTARC1 levels, helping assess therapeutic opportunities and potential safety considerations for MTARC1 inhibition. To assess potential broader health implications relevant to target development, we performed a phenome-wide MR study across. These analyses confirmed an effect of higher MTARC1 levels on MASLD and related phenotypes and an opposite effect on gout. These findings should be replicated in randomized controlled trials of MTARC1 inhibition in MASLD.

### Importance of assessing liver protein levels beyond expression quantitative trait loci

Although pQTLs and eQTLs frequently overlap in some tissues (e.g., ∼75% overlap in brain), liver exhibits considerably lower concordance (∼50% overlap) [54]. Many eQTLs do not translate into protein changes, highlighting post-transcriptional regulation or that roughly half of liver pQTLs act through post transcriptional or post translational mechanisms rather than mRNA abundance. On a functionality perspective, pQTLs tend to be more directly linked to diseases, and are often variants that alter protein structure or stability (non-synonymous) like the MTARC1 variant rs2642438. Interestingly, this variant was not a liver eQTL. In other loci, such as GCKR, the direction of effect at the transcript level (eQTL) and at the protein level (pQTL) was discordant, highlighting that genetic effects on mRNA abundance may not provide valuable information on disease biology and that mechanistic inference based on eQTLs alone can be incomplete and, in some settings, misleading with respect to the functional molecular phenotype. Integrating protein levels on top of gene expression levels therefore provides a more complete picture of MASLD genetic and molecular architecture.

### Biomarkers rather than potential targets

A key insight from comparing proteomic DEPs to genetic evidence is that some of the strongest tissue-level changes likely reflect adaptive responses to lipid stress and injury rather than causal drivers. Among the proteins whose relative abundance were significantly higher in MASL than control condition, PLIN1 and PLIN2 demonstrated the largest fold-changes, with MASLD participants exhibiting 24% and 47% higher hepatic concentrations, respectively. Both proteins belong to the perilipin family and localize to lipid droplets, where they regulate lipid storage and lipolysis. However, these candidates did not emerge as causal mediators in MR/colocalization analyses (Suplementary Table 3). Similarly, LGALS1 and DUSP23 may show state-dependent changes consistent with inflammatory signaling or stress-response pathways, yet the genetic evidence did not support a primary causal role (Suplementary Table 3). Such discordance provides mechanistic stratification: these proteins may be important for disease expression and progression, but their upregulation may be reactive, compensatory, or downstream of primary metabolic perturbations driven by inherited risk pathways. This distinction is critical for therapeutic translation. Causal proteins may represent upstream levers capable of modifying disease susceptibility, whereas adaptive proteins may still be biologically important but could be riskier or less effective to target directly, particularly if they represent protective responses or general stress programs. However, they could represent specific biomarkers for MASLD. To date, no MASLD specific clinical trials directly evaluate PLIN1 as a diagnostic or therapeutic endpoint. Clinical studies have demonstrated that circulating PLIN2 levels in monocytes and plasma strongly discriminate MASH and advanced fibrosis, outperforming existing non invasive scores [55, 56]. A large, ongoing interventional validation study (TEIRESEAS; NCT07502755) is currently assessing PLIN2 (alone and in combination with RAB14) for diagnosis and therapeutic monitoring of biopsy confirmed NASH with fibrosis in >800 participants. This builds on prior multicenter trials (BRAVES, LIBRA) showing high accuracy (AUC >0.9) for NASH and fibrosis detection using PLIN2 based liquid biopsy approaches [57]. While DUSP23 has emerged as a prognostic marker in solid tumors, including liver cancer datasets, no clinical trials currently evaluate DUSP23 in MASLD [58, 59]. Public expression atlases show measurable hepatic expression, but its role in steatosis, inflammation, or fibrosis remains poorly defined, and there is no therapeutic or biomarker driven MASLD program involving DUSP23 currently.

### Limitations and future directions

Several limitations should be considered. First, the proteomic comparisons are cross-sectional and therefore cannot definitively establish temporal ordering of molecular changes across disease stages. Second, liver tissue reflects a mixture of cell types; differential abundance may arise from hepatocyte-intrinsic regulation, immune infiltration, or changes in stromal composition during fibrosis. Third, while MR and colocalization strengthen causal inference, they rely on assumptions about instrument validity, absence of horizontal pleiotropy, and adequate power.

Finally, functional experiments will be necessary to validate whether genetically supported proteins such as MTARC1, HSD17B13, ERLIN1, and others directly modulate hepatocyte lipid handling, inflammatory signaling, or fibrogenesis in relevant cellular and in vivo models. Future work should focus on cell-type-resolved proteomics or spatial profiling to localize DEPs to specific hepatic compartments; longitudinal sampling to connect early metabolic changes to subsequent fibrosis; mechanistic perturbation studies for genetically supported proteins; and evaluation of whether state-dependent DEPs (e.g., PLIN1/PLIN2) improve risk stratification, staging, or treatment response monitoring when combined with clinical and imaging measures.

## Conclusions

Integrating liver proteomic with gene expression levels and human genetics provides a powerful framework to understand of how genetic variations influence MASLD across molecular layers. While proteomics revealed extensive metabolic and immunometabolic remodeling in MASLD and highlighted inflammatory and structural changes in MASH, overlap with genetic evidence was limited, indicating that proteomic disease signatures may reflect consequence and adaptation, rather than causal drivers. By integrating DEP, pQTL-based MR, and colocalization, we prioritized a set of concordant proteins (MTTP, TM6SF2, ERLIN1, HSD17B13, MTARC1, and SERPINA1) as high-confidence candidates for causal involvement and therapeutic exploration. Overall, this integrative analysis enabled the distinction between causal mechanisms from reactive signatures providing a rational strategy to prioritize of targets for MASLD intervention.

## Acknowledgements

The authors would like to thank all study participants as well as all investigators of the studies that were used throughout the course of this investigation. We acknowledge the invaluable collaboration of the surgery team, bariatric surgeons, and Quebec Obesity Biobank staff and would like to thank all study participants from the *Institut universitaire de cardiologie et de pneumologie de Québec* (IUCPQ).

## Funding

This study was supported by the Canadian Institutes of Health Research (CIHR) and the Fondation IUCPQ. The funders had no role in the design of the study; in the collection, analyses, or interpretation of data; in the writing of the manuscript, or in the decision to publish results. ÉG and LJR hold a Doctoral Research Award from the CIHR. BJA holds a Senior Scholar Award from the FRQS.

## Competing interests

BJA is a consultant for Novartis, MSD, Eli Lilly, and Silence Therapeutics and has received research contracts from Pfizer, Eli Lilly and Silence Therapeutics. The remaining authors disclose no conflicts.

## Notes

### Author Declarations

The study was conducted in accordance with the Declaration of Helsinki and approved by the Institutional Review Board (Ethics Committee) of Institut universitaire de cardiologie et de pneumologie de Quebec-Universite Laval (IUCPQ-UL) (approval number 2021-3656; date of approval: 17 June 2021). For the other publicly available GWAS summary statistics, all participants provided informed consent, and study protocols were approved by their respective local ethical committee.

